# High prevalence of low high-density lipoprotein cholesterol and insulin resistance among children and adolescents living with HIV in Uganda: Harbinger for metabolic syndrome?

**DOI:** 10.1101/2022.12.02.22282969

**Authors:** Grace Kisitu, Veronika Shabanova, Fairuzi Naiga, Mary Nakagwa, Adeodata R Kekitiinwa, Peter J. Elyanu, Elijah Paintsil

**Author notes:** **Correspondence to:** Elijah Paintsil, MD., Department of Pediatric, Yale School of Medicine, 464 Congress Avenue, New Haven, Connecticut 06520, USA.; Or Peter J. Elyanu, MBChB, M.Med, PhD., Baylor College of Medicine Children’s Foundation-Uganda, Block 5 Mulago Hospital, P.O Box 72052, Kampala, Uganda. GK and VS contributed equally.

## Abstract

**Background:** Antiretroviral Therapy-associated adverse effects and comorbidities are still pervasive in people living with HIV, especially metabolic syndrome (MetS), which is on the rise and occurring at early age. However, there is paucity of data on MetS in children and adolescents living with HIV (CALWH), particularly in sub-Saharan Africa. We investigated the age-dependent prevalence of components of MetS in this population.

**Methods:** A cross-sectional pilot study of CALWH treated at the Baylor Uganda Clinical Centre of Excellence in Kampala, Uganda. Using stratified by age group and sex random sampling, participants were recruited from May to August 2021. At enrollment, we collected data on participant demographics, anthropometric measurements, HIV disease characteristics, and past medical history and obtained blood for fasting levels of glucose, insulin, triglycerides, total cholesterol, and high-density lipoprotein (HDL) cholesterol. The primary outcome of MetS was defined by both the International Diabetes Federation (IDF) and Adult Treatment Panel (ATPIII) criteria. We estimated the prevalence of MetS and its components for all participants and by the stratification factors.

**Results:** We enrolled 90 children and adolescents: <10 y/o (N=30), 10 to <16 y/o (N=30), and ≥16 (N=30) y/o. Fifty-one percent were females. The prevalence of MetS was 1.11% (1 of 90) using either IDF or ATPIII criteria for all participants, and 3.33% for ≥16-year group. Over 55% of participants had ≥1 IDF component, with 47% having low HDL cholesterol; 14% of participants had early insulin resistance using the HOMA index. The proportion of early insulin resistance was 6.67%, 23.33%, and 13.33% for the three age groups, respectively. Two participants (6.67%) in the 10 to <16 years group had significant insulin resistance. For every 1-year increase in age, HOMA index increased by 0.04 (95% Confidence Interval 0.01, 0.08), p=0.02.

**Conclusions:** The high prevalence of components of MetS, particularly low HDL and early insulin resistance, are of concern. With increasing survival of CALWH into adulthood and increased lifetime exposure to ART, the frequency of MetS in this population may rise, increasing the lifetime risk for associated health problems, such as type 2 diabetes, myocardial infarction, stroke, and nonalcoholic fatty liver disease.

## Introduction

Although contemporary combination antiretroviral therapy (ART) has reduced HIV-associated morbidity and mortality drastically, ART-associated adverse effects and comorbidities are still pervasive in people living with HIV (PLWH)^1-3^. Notable among these is metabolic syndrome (MetS), which appears to be in the final common pathway of most non-communicable diseases. In adults, MetS is defined by at least three of five components, i.e., impaired fasting glucose or diabetes, hypertension, central obesity (increased waist circumference), elevated triglycerides, or reduced high-density lipoprotein (HDL) cholesterol^4^. Furthermore, MetS increases risk for many health problems such as type 2 diabetes, myocardial infarction, stroke, and nonalcoholic fatty liver disease^5,6^. The prevalence of MetS among PLWH is high, ranging from 17% to 45% and it is occurring at an earlier age compared to HIV-uninfected populations^7-9^. Interestingly, the etiopathogenesis of MetS in HIV-infected is complex and not well understood; HIV infection per se, exposure to ART, and traditional cardiovascular (CDV) risk factors (e.g., genetic factors, nutrition, physical activity) contribute to the etiopathogenesis.

In 2007, the International Diabetes Federation (IDF) published the first consensus definition of MetS in children and adolescents: (1) for children 6 years to <10 years old, obesity (defined as ≥90^th^ percentile of waist circumference); (2) for age 10 to <16 years, obesity (defined as waist circumference ≥90^th^ percentile), and the adult criteria for abnormal triglycerides, HDL cholesterol, blood pressure, and glucose; and (3) for youth ≥16 years of age adult criteria are used^10^. A systematic review of all pediatric studies (age ranging from <7 to 19 years) on MetS published from 2003-2012 reported a median prevalence of 3.3% (range, 0-19.2%) in the general pediatric population, 11.9% (range, 2.8-29.3%) in overweight children, and 29.2% (range, 10-66%) in obese children^11^. Unfortunately, there is paucity of studies on the true prevalence of MetS (i.e., meeting the full diagnostic criteria) in children and adolescent living with HIV (CALWH). Most MetS studies in HIV+ children report only the incidence of separate components of MetS, ranging from 20% to 86%^12-24^. A recent cross-sectional study from Spain reported the incidence of MetS in HIV+ children (a median age of 12.5 years (interquartile range: 9.4–16.2 years) to be 7.4%^25^ using the Third Report of the National Cholesterol Education Program Expert Panel on Detection, Evaluation, and Treatment of High Blood Cholesterol in Adults (NCEP-ATP III) ^26^ criteria.

With increasing survival of perinatally HIV+ children on ART into adulthood and with prolonged exposure to ART, the incidence of MetS in this population may rise. Also, it is not known whether the high prevalence of individual components of MetS is transient or may evolve into full-blown MetS over time. Furthermore, it is unclear whether MetS in children increases the lifetime risk of cardiovascular diseases as in adults^27,28^. There is an urgent need to find out the true prevalence of MetS in CALWH in sub-Saharan Africa, home to over 90% of children and adolescents living with HIV, because the effects of MetS in this population may be more devastating than in adults due to the long and cumulative exposure to ART. In this study, we investigated the age-dependent prevalence of MetS and its components in HIV+ children and adolescents receiving care at the Baylor-Uganda Clinical Centre of Excellence (COE) in Kampala, Uganda.

## Methods

### Study site, population, and design

The study was conducted at the Baylor-Uganda Clinical COE in Kampala, Uganda. The Baylor-Uganda Clinical COE is the largest provider of children and adolescent HIV care in Uganda. The clinic serves about 5000 children and adolescents living with HIV aged <20 years, of whom 99% are on ART. Inclusion criteria: HIV infected children and adolescents aged 6-19 years of age, on combination ART comprised of at least three drugs from two different classes, and who provided consent and assent to participate in the study. Exclusion criteria: HIV+ individual <6 or >19 years of age; history of hepatitis B or C infection, diabetes, thyroid diseases, renal disease, cardiac disease; severe malnutrition; and history of mitochondrial disorder or other conditions known to affect mitochondrial function or lactate levels (e.g., Acute infection, malignancy, and ischemic conditions).

The study was approved by ethics committees at School of Biomedical Sciences Higher Degree, Makerere University, Uganda, Uganda National Counsel for Science and Technology, and Yale School of Medicine, and informed consents or assents for participation was obtained from all patients and parents or legal guardians, as appropriate.

Sample size estimation. The true prevalence of MetS in children (HIV+ and HIV-) in sub-Saharan Africa is not known so we had no data for the sample size calculations. Therefore, the study was an exploratory pilot study to obtain preliminary estimates of the true prevalence of MetS and its components. We enrolled a total of 90 CALWH and based on the age categorization for MetS definition: 6 to <10 years (N=30), 10 to <16 years (N=30), and ≥16 years (N=30). Given that the available estimate of MetS prevalence among children not infected with HIV is 0.033 (range 0.0-0.192)^11^, with 30 children in each age stratum, the 95% Confidence Interval (95% CI) around the estimate of 0.033 would be 0.001 to 0.17; thus, with a total sample of 90 children, the 95% CI would be 0.01 to 0.09^29,30^.

### Study procedures and measures

#### Procedures

Using the eligibility criteria, potential study participants were identified from the Baylor-Uganda electronic medical record database. We used stratified random sampling to select participants. The strata included three age bands: 6 to <10, 10 to <16, and 16 - 19 years. Within each stratum, systematic random sampling based on the weekly clinic schedule patient roster was conducted to identify which participants to contact to participate in the study. A list of patients to be contacted and their matched alternates (in case of a no-show or a refusal) was used by the study nurse to phone caregivers and legal guardians, in order to provide study information. Those who were interested were invited to the clinic for study screening and consenting. Upon arrival of potential study participants to the clinic, the study nurse explained the purpose of the study and procedures to the caregiver-child dyad, answered their questions, and obtained informed consent and assent accordingly. While the interviews and physical examinations were done on day of enrollment, blood samples were taken the following day after the participant had fasted overnight, for at least 8 hours after their last meal the previous night. Participants who declined to participate were replaced by sampling other participants from the age-specific sampling frame.

#### Study survey

At enrollment, the study physician examined participants and collected data using a study survey. The survey included questions on demographic characteristics, other past medical history, smoking, exercise and activity level, dieting, and general lifestyle questions. The participant’s medical record was extracted for date of HIV diagnosis, mode of HIV transmission, medication history (past and current ART regimens, non-HIV medication use), past medical history, clinical and immunologic staging of their disease, HIV RNA Viral load, and CD4+ T-cell count.

#### Anthropometric measurements

A study nurse used standardized techniques for vital signs and anthropometric measurements. Weights and heights were taken with the participants in light clothing and barefooted. Weights and heights were measured to the nearest 0.1 kg and 0.1 cm, respectively. Mid-upper arm and waist circumferences were measured to the nearest 0.1 cm using a measuring tape. Waist circumference (WC) was measured at the midpoint between the lowest rib lateral and iliac crest. Body Mass Index (BMI) was calculated as weight in kg divided by height in meters squared. Blood pressure was measured using sphygmomanometer with the age-appropriate size cuff in the right arm in the sitting position. An average of 2 readings taken 5 minutes apart were recorded for analysis.

#### Biochemical parameters

Fasting glucose, insulin, triglycerides, total cholesterol, and high-density lipoprotein (HDL) cholesterol tests were analyzed by standard laboratory methods at the Baylor-Uganda Clinical COE Laboratory. The Baylor-Uganda Clinical COE laboratory is certified as a College of American Pathologists (CAP) laboratory since May 2013.

#### Definitions

We used both International Diabetes Federation (IDF) and Adult Treatment Panel (ATP) III definitions for MetS. The IDF criteria is based on age as described above^10^ and the presence of central obesity plus at least two of four other factors. ATP III definition of MetS is abnormal values for at least 3 of the 5 following: Triglyceride≥1.7 mmol/L; HDL <1.03 mmol/L for males and <1.3 mmol/L for females; waist circumference ≥90^th^ percentile for age; fasting glucose ≥5.6 mmol/L; and blood pressure ≥90^th^ percentile for age, sex, and height^31^. The homeostatic model assessment of insulin resistance (HOMA-IR) was calculated as the product of insulin (mIU/L) and glucose (mmol/L) divided by 22.5^32^.

### Statistical analysis

Participant demographics and characteristics were summarized using mean (standard deviation, SD) for continuous data and counts (n, %) for categorical data. These were compared among the three age groups using the Kruskal-Wallis test with post-hoc pair-wise age group comparisons based on the Wilcoxon Rank Sum test for continuous variables, and using the Chi-square or Fisher’s Exact test for the categorical variables. The prevalence of MetS in children using both IDF and NCEP-ATPIII definitions was reported with a surrounding bootstrapped 95% CI (using 5000 bootstrap samples). We estimated the prevalence for all children and adolescents, as well as stratified by the three age groups. We also estimated and compared the frequency of the individual components of MetS diagnosis among the three age groups, using the Armitage trend test. No adjustments of the significance level for multiple comparisons were done due to the pilot nature of the study, the purpose of which was to obtain preliminary estimates of relevant effect sizes which could be used to design a larger study.

## Results

### Characteristics of study participants

We enrolled 90 children and adolescents stratified into three age bands according to IDF MetS definition— <10 (N=30), 10 to <16 (N=30), and ≥16 (N=30) years— from Baylor-Uganda Clinical COE between May and August 2021. The demographic and anthropometric characteristics of study participants are shown in Table 1. Fifty one percent were females and the mean (SD) age for the entire study population was 12.96 (4.14) years. The proportions of participants at the different educational levels were commensurate with the age groups. The differences in Tanner staging, height, weight, and mid upper arm circumference (MUAC) were as expected for the age groups. Over 75% of the children and adolescents were in the care of their mother and/or father. The mean age of caregivers was 40.08 (9.83) years, with the caregivers of children <10 years of age much younger, 35.69 (8.50) years.

**Table 1:**
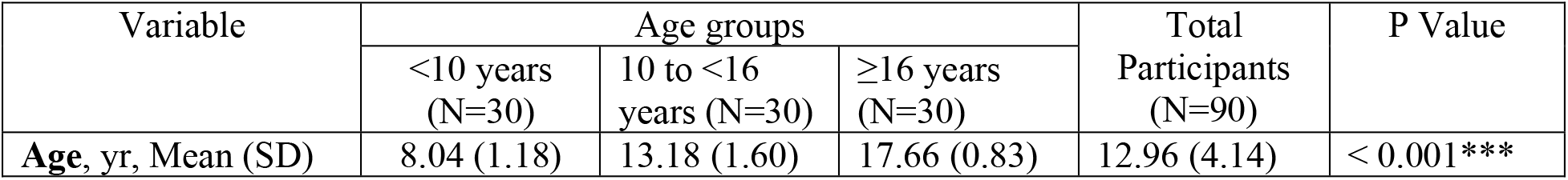
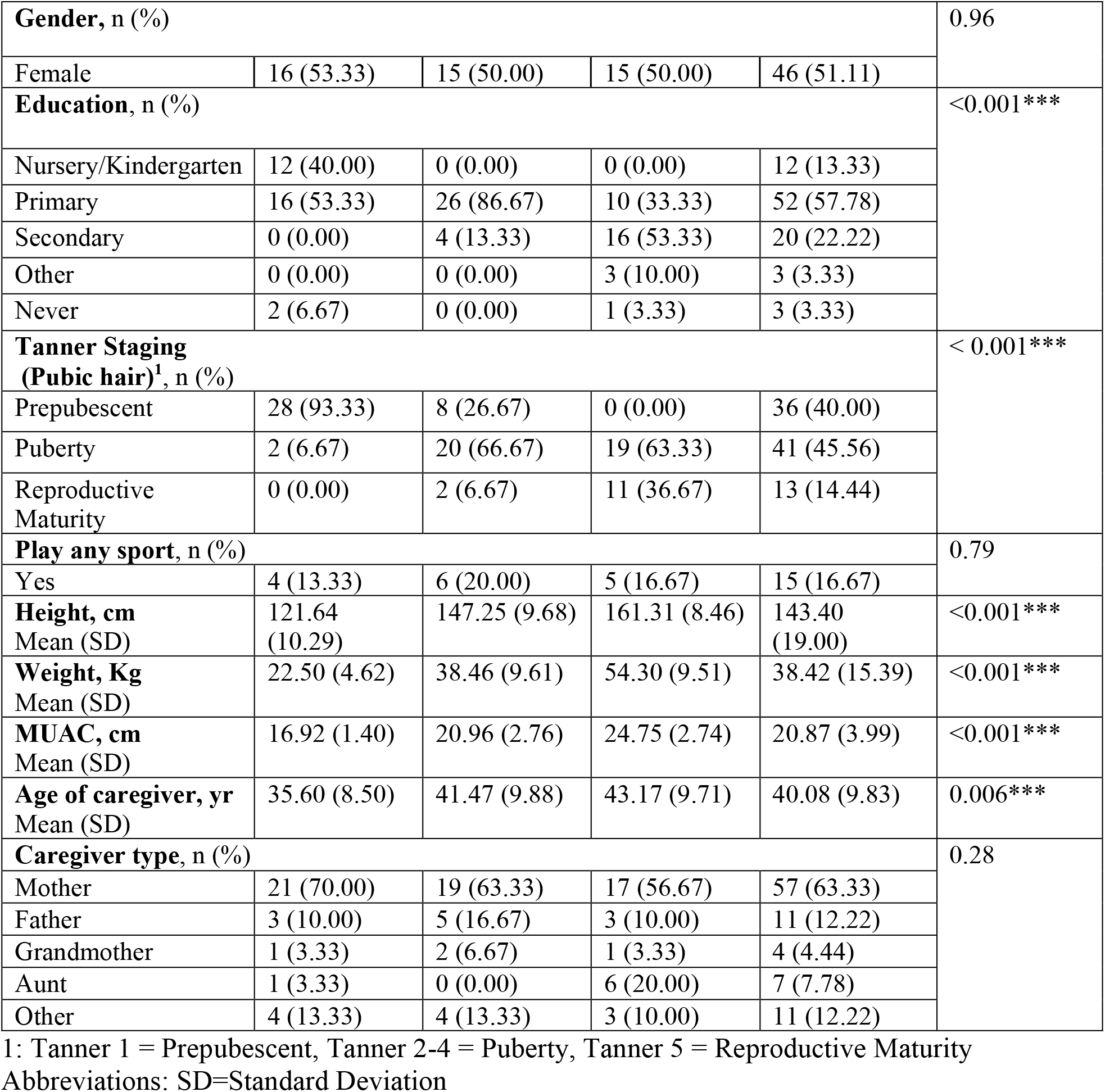
Demographic and anthropometric characteristics of study participants, age stratified

| Variable | Age groups |  |  | Total<br>Participants<br>(N=90) | P Value |
| --- | --- | --- | --- | --- | --- |
| | <10 years<br>(N=30) | 10 to <16<br>years (N=30) | ≥16 years<br>(N=30) | | |
| **Age, yr, Mean (SD)** | 8.04 (1.18) | 13.18 (1.60) | 17.66 (0.83) | 12.96 (4.14) | < 0.001*** |
| **Gender, n (%)** |  |  |  |  | 0.96 |
| Female | 16 (53.33) | 15 (50.00) | 15 (50.00) | 46 (51.11) |  |
| **Education, n (%)** |  |  |  |  | <0.001*** |
| Nursery/Kindergarten | 12 (40.00) | 0 (0.00) | 0 (0.00) | 12 (13.33) |  |
| Primary | 16 (53.33) | 26 (86.67) | 10 (33.33) | 52 (57.78) |  |
| Secondary | 0 (0.00) | 4 (13.33) | 16 (53.33) | 20 (22.22) |  |
| Other | 0 (0.00) | 0 (0.00) | 3 (10.00) | 3 (3.33) |  |
| Never | 2 (6.67) | 0 (0.00) | 1 (3.33) | 3 (3.33) |  |
| **Tanner Staging (Pubic hair)<sup>1</sup>, n (%)** |  |  |  |  | < 0.001*** |
| Prepubescent | 28 (93.33) | 8 (26.67) | 0 (0.00) | 36 (40.00) |  |
| Puberty | 2 (6.67) | 20 (66.67) | 19 (63.33) | 41 (45.56) |  |
| Reproductive Maturity | 0 (0.00) | 2 (6.67) | 11 (36.67) | 13 (14.44) |  |
| **Play any sport, n (%)** |  |  |  |  | 0.79 |
| Yes | 4 (13.33) | 6 (20.00) | 5 (16.67) | 15 (16.67) |  |
| **Height, cm**<br>Mean (SD) | 121.64<br>(10.29) | 147.25 (9.68) | 161.31 (8.46) | 143.40<br>(19.00) | <0.001*** |
| **Weight, Kg**<br>Mean (SD) | 22.50 (4.62) | 38.46 (9.61) | 54.30 (9.51) | 38.42 (15.39) | <0.001*** |
| **MUAC, cm**<br>Mean (SD) | 16.92 (1.40) | 20.96 (2.76) | 24.75 (2.74) | 20.87 (3.99) | <0.001*** |
| **Age of caregiver, yr**<br>Mean (SD) | 35.60 (8.50) | 41.47 (9.88) | 43.17 (9.71) | 40.08 (9.83) | 0.006*** |
| **Caregiver type, n (%)** |  |  |  |  | 0.28 |
| Mother | 21 (70.00) | 19 (63.33) | 17 (56.67) | 57 (63.33) |  |
| Father | 3 (10.00) | 5 (16.67) | 3 (10.00) | 11 (12.22) |  |
| Grandmother | 1 (3.33) | 2 (6.67) | 1 (3.33) | 4 (4.44) |  |
| Aunt | 1 (3.33) | 0 (0.00) | 6 (20.00) | 7 (7.78) |  |
| Other | 4 (13.33) | 4 (13.33) | 3 (10.00) | 11 (12.22) |  |

1: Tanner 1 = Prepubescent, Tanner 2-4 = Puberty, Tanner 5 = Reproductive Maturity Abbreviations: SD=Standard Deviation

### HIV disease characteristics of participants (Table 2)

The mode of HIV transmission in all the participants was through mother-to-child-transmission, MTCT. The mean (SD) duration of HIV diagnosis was 6.12 (2.42), 11.07 (1.94), and 10.30 (3.84) for <10, 10 to <16, and ≥16 year groups, respectively (Table 2). In a decreasing order as expected, the mean (SD) CD4 count was 1330.87 (993.39), 1060.22 (893.87), and 798.74 (600.28) cells/mm^3^ for <10, 10 to <16, and ≥16 year groups, respectively. Participants in the <10 year group tended to have a higher proportion with WHO stages 4 and 5. Over 90% of participants had experienced changes in their ART regimen. About 90% of all participants had at least 3 changes in their ART regimen over a mean duration of ART of 9.42 (3.73) years.

**Table 2:**
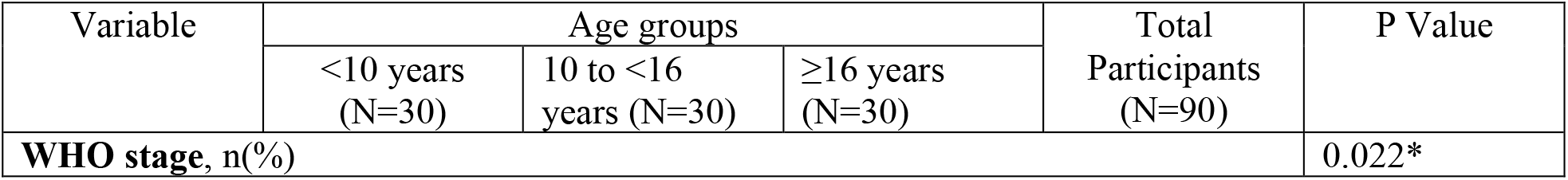
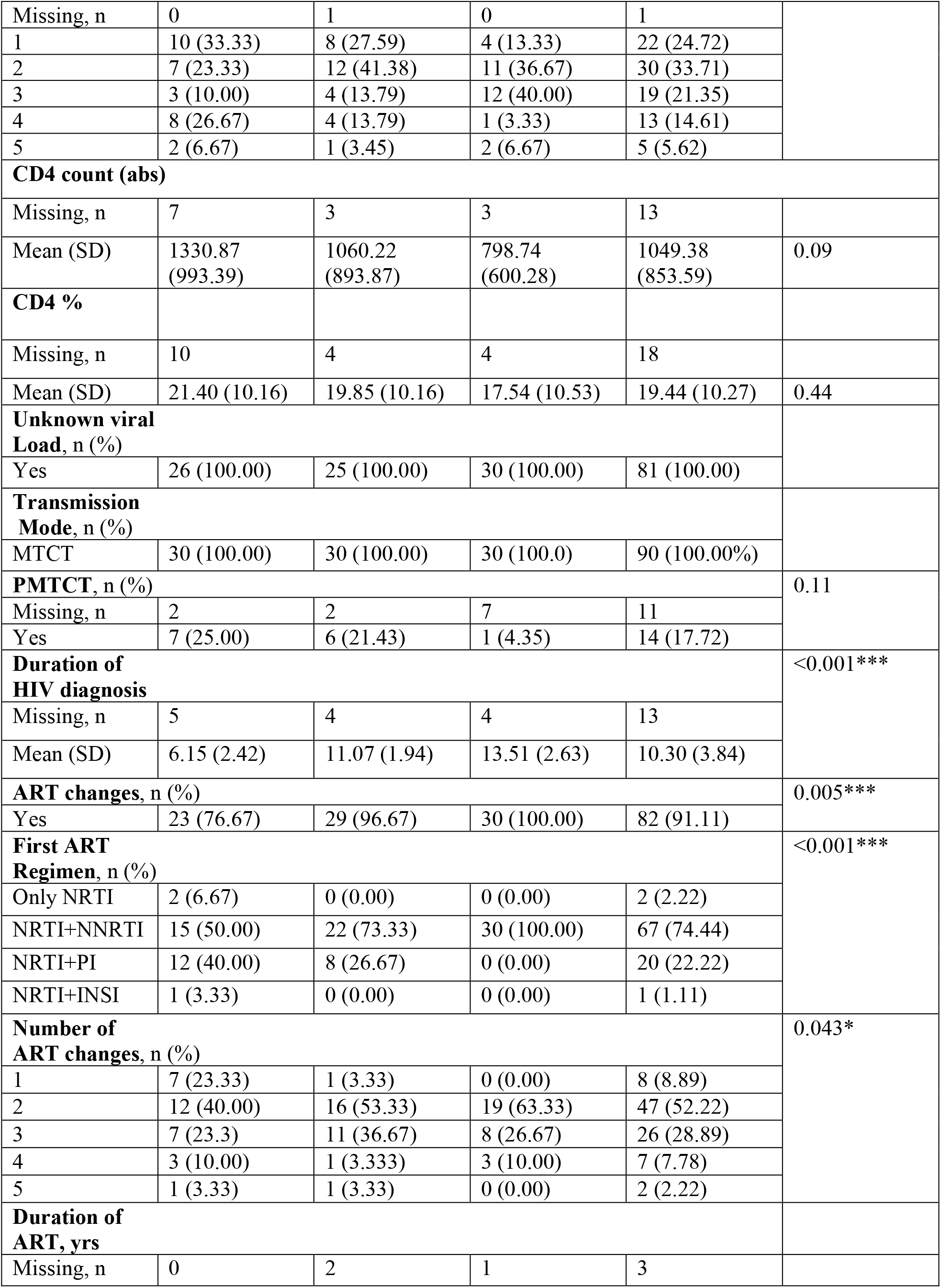
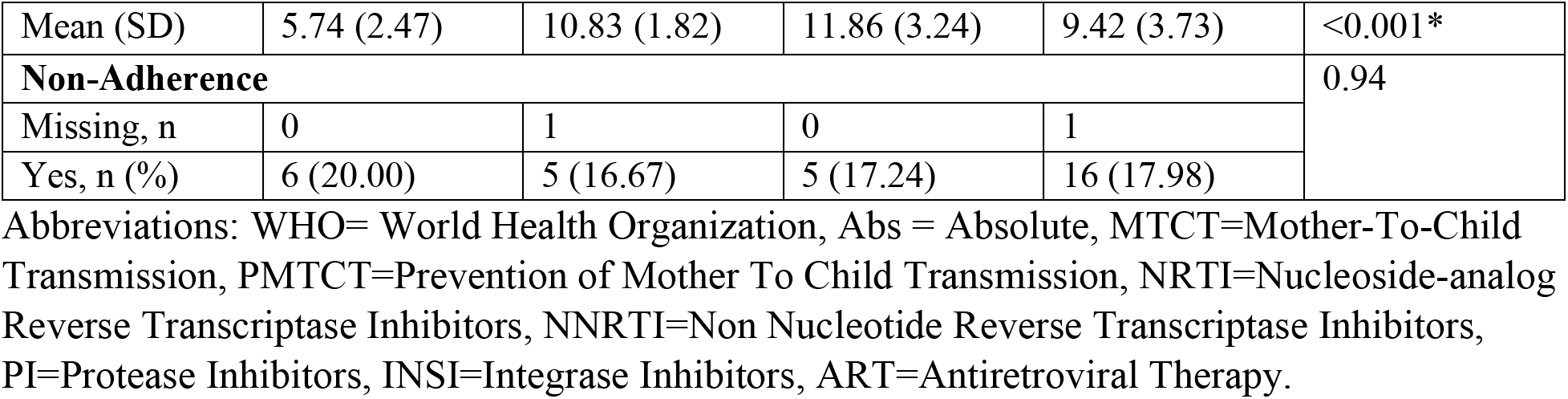
Characteristics of HIV disease of participants, age stratified

### Prevalence of components of MetS criteria (Table 3)

There were growth-related differences across the age groups, as expected, with regard to systolic BP, diastolic BP, and waist circumference. Total cholesterol and LDL cholesterol were similar across the age groups, but mean HDL cholesterol decreased from younger to older age groups. Although the mean triglyceride levels for all ages were within the normal range (≤1.7 mmol/L), younger children had lower levels as compared to older children. Fasting blood sugar level was within normal for all ages, however, fasting insulin level tended to increase with age. Mean HOMA index was the highest in the older age groups as illustrated in the boxplot (Figure 1A). We found that for every 1-year increase in age, HOMA index increased by 0.04 (95%CI 0.01, 0.08), p=0.015 (Figure 1B). Fourteen percent (14%) of all participants, with higher prevalence among the older age groups, showed early insulin resistance as defined by HOMA index >1.9^33^. Two participants (6.67%) in the 10 to 16 years group were classified as having significant insulin resistance (HOMA of >2.9) (Table 3). BMI increased across the age groups (Figure 2).

**Table 3:** Components of MetS diagnosis, age stratified

| Variable | Age groups |  |  | Total Participants (N=90) | P Value |
| --- | --- | --- | --- | --- | --- |
|  | <10 years (N=30) | 10 to <16 years (N=30) | ≥16 years (N=30) |  |  |
| **Systolic BP**<br>Mean (SD) | 102.57 (9.67) | 105.60 (9.75) | 110.83 (10.21) | 106.33 (10.35) | 0.006** |
| **Diastolic BP**<br>Mean (SD) | 63.57 (7.89) | 67.93 (5.95) | 71.50 (7.30) | 67.67 (7.74) | <0.001*** |
| **Waist Circumference, cm**<br>Mean (SD) | 54.67 (4.02) | 62.63 (6.02) | 71.09 (8.48) | 62.80 (9.27) | <0.001*** |
| **Total Cholesterol, mmol/L**<br>Mean (SD) | 3.80 (0.73) | 3.82 (0.83) | 3.53 (0.99) | 3.72 (0.86) | 0.16 |
| **Triglycerides, mmol/L**<br>Mean (SD) | 1.25 (0.53) | 1.12 (0.46) | 0.84 (0.36) | 1.07 (0.48) | <0.001 |
| **HDL Cholesterol, mmol/L**<br>Mean (SD) | 1.22 (0.39) | 1.18 (0.19) | 1.10 (0.22) | 1.16 (0.28) | 0.045 |
| **LDL Cholesterol, mmol/L** | 2.36 (0.57) | 2.46 (0.75) | 2.39 (0.82) | 2.41 (0.71) | 0.86 |
| **Fasting Blood Sugar- Mean (SD)** | 4.26 (0.45) | 4.56 (0.47) | 4.28 (0.66) | 4.34 (0.55) | 0.03* |
| **Fasting Insulin, Mean (SD)** | 3.75 (1.95) | 7.06 (3.86) | 6.17 (2.98) | 5.66 (3.31) | < 0.001*** |
| **HOMA-IR Mean (SD)** | 0.73 (0.43) | 1.45 (0.83) | 1.15 (0.55) | 1.11 (0.69) | < 0.001*** |
| **Early IR, n (%)** |  |  |  |  |  |
| ≤1.9 | 28 (93.33) | 23 (76.67) | 26 (86.67) | 77 (85.56) | 0.10 |
| >1.9 | 2 (6.67) | 7 (23.33) | 4 (13.13) | 13 (14.44%) |  |
| **Significant IR** |  |  |  |  |  |
| ≤2.9 | 30 (100.00) | 28 (93.33) | 30 (100.00) | 88 (97.78) | 0.33 |
| >2.9 | 0 (0.00) | 2 (6.67) | 0 (0.00) | 2 (2.22) |  |

### Prevalence of MetS components and full diagnostic criteria for IDF and ATPIII (Table 4)

The prevalence of MetS was 1.11% (1 of 90, 95%CI 0.1%, 3.3%) using either IDF or ATPIII criteria for all participants, and 3.33% (95%CI 0.1%, 10.0%) for ≥16-year group. About 11% and 47% of participants had high triglyceride levels and low HDL cholesterol levels, respectively. While the younger age group tended to have high triglyceride levels, proportion with low HDL cholesterol increased with age; 57% of ≥16 age group had low HDL cholesterol level. About 56% of participants had ≥1 (1 to 3) components of ATPIII MetS diagnostic criteria, while over 55% of participants had ≥1 (1 to 2) components of IDF MetS diagnostic criteria.

**Table 4:** Participants meeting individual components and full MetS criteria

| Variable | Age groups |  |  | P Value |
| --- | --- | --- | --- | --- |

|  | <10 years<br>(N=30) | 10 to <16<br>years (N=30) | ≥16 years<br>(N=30) | Total<br>Participants<br>(N=90) |  |
| --- | --- | --- | --- | --- | --- |
| **Triglycerides, n (%)** |  |  |  |  | 0.15 |
| Yes | 6 (20.00) | 3 (10.00) | 1 (3.33) | 10 (11.11) |  |
| **HDL, n (%)** |  |  |  |  | 0.10 |
| Yes | 12 (40.00) | 13 (43.33) | 17 (56.67) | 42 (46.67) |  |
| **Fasting Blood**<br>**Sugar, n (%)** |  |  |  |  |  |
| Yes | 0 (0.00) | 0 (0.00) | 0 (0.00) | 0 (0.00) |  |
| **Number of**<br>**criteria for**<br>**ATPIII, n (%)** |  |  |  |  | 0.10 |
| 0 | 15 (50.00) | 14 (46.67) | 11 (36.67) | 40 (44.44) |  |
| 1 | 12 (40.00) | 15 (50.00) | 17 (56.67) | 44 (48.89) |  |
| 2 | 3 (10.00) | 1 (3.33) | 1 (3.33) | 5 (5.56) |  |
| 3 | 0 (0.00) | 0 (0.00) | 1 (3.33) | 1 (1.11) |  |
| **Full ATPIII**<br>**Criteria, n (%)** |  |  |  |  | 1.00 |
| Yes | 0 (0.00) | 0 (0.00) | 1 (3.33) | 1 (1.11) |  |
| **Number of**<br>**criterial for IDF** |  |  |  |  | 0.10 |
| 0 | 15 (50.00) | 14 (46.67) | 11 (36.67) | 40 (44.44) |  |
| 1 | 12 (40.00) | 15 (50.00) | 17 (96.67) | 44 (48.89) |  |
| 2 | 3 (10.00) | 1 (3.33) | 2 (6.67) | 6 (6.67) |  |
| **Full IDF**<br>**Criteria, n (%)** |  |  |  |  | 1.00 |
| Yes | 0 (0.00) | 0 (0.00) | 1 (3.33) | 1 (1.11) |  |

Abbreviations: ATPIII= Adult Treatment Panel (ATPIII); IDF=International Diabetes

## Discussion

In this exploratory pilot cross-sectional study, we sought to investigate the age-dependent prevalence of components of MetS in children and adolescents living with HIV (CALWH) receiving care at Baylor-Uganda Clinical COE in Kampala. The prevalence of full-blown MetS was 1.11% (1 of 90) using either IDF or ATPIII criteria for all participants, and 3.33% for ≥16-year group. To our surprise, over half of the participants had ≥1 components of either IDF or ATPIII diagnostic criteria; 47% had low HDL cholesterol and 14% of participants had early insulin resistance using the HOMA index: 6.67%, 23.33%, and 13.33% for the three age groups, respectively. Of note, none of the participants met the criteria for obesity based on their age-sex. Our study raises two important questions: (1) Could the observed underweight and/or normal BMI have resulted in under diagnosis of MetS, particularly using the IDF criteria which requires the presence of central obesity plus at least two of four other factors? (2) Is the high prevalence of MetS components a transient phenomenon or harbinger for full-blown MetS with time?

There are few studies in CALWH on the prevalence of MetS meeting the full diagnostic criteria. Our findings of prevalence of 1.1% and 3.3% in all participants and ≥16 years group, respectively, are consistent the prevalence of MetS in the general pediatric population reported in a systematic review of studies published from 2003-2012; a median prevalence of 3.3% (range, 0-19.2%) ^11^. Moreover, Espiau et al. reports a prevalence of 1.97% and 5.92% using IDF and NCEP-ATP III criteria, respectively, in CALWH in Spain^34^. There are no reports from sub-Saharan Africa on either true prevalence or incidence of Mets in CALWH. Could this be due to non-reporting of negative studies and/or inconsistencies in the diagnostic criteria of MetS? For instance, the IDF criteria, the accepted consensus definition in pediatrics, requires that and individual has central obesity as a minimum criterion. In CALWH, central obesity may not be present even in those with lipodystrophy syndrome^13,24^. Even among adults, central obesity is less prevalent in PLWH compared to non-HIV-infected population^35^. Moreover, in sub-Sharan Africa, home to over two-thirds of CALWH globally, stunting and undernutrition are still pervasive^36^ and may affect any MetS diagnostic definition dependent on weight and/or body habitus. These children are experiencing pubertal changes which may further confound MetS diagnosis^37,38^. Taken together, there is a need for validation of MetS diagnostics criteria and systematic longitudinal study of MetS in CALWH.

Most MetS studies in HIV+ children report only the incidence of components of MetS ranging from 20% to 86%^12-23^. Over half of our cohort had ≥1 components of either IDF or ATP III diagnostic criteria, with high triglycerides (11%) and low HDL cholesterol levels (47%) being the most prevalent components of MetS. These findings are consistent with previous findings in HIV-infected children^34,39^ and adults^40^. Limited studies from sub-Saharan Africa have reported a much lower prevalence of HDL (6% to 17%) and triglyceride (6% to 9%), but these studies were in prepubertal HIV+ children^17,41,42^. In a study from 12 clinical sites in Latin America of HIV+ children with a mean age of 6.6 years, the prevalence of low HDL and triglyceride was 20% and 44%, respectively^43^. In a systematic review of articles published from 2008 to 2014 on MetS in adults living with HIV, Naidu et al. reported that the mean prevalence of hypertriglyceridemia in studies from Africa was 24.9% and low HDL cholesterol was the most frequent (50.1%) reported component of MetS from developing countries^44^. In a pediatric study from Spain, low HDL cholesterol level was the most prevalent (21.05%)^34^. In a study in Thailand of youth with HIV aged 15 to <25 years, the prevalence of low HDL was 47%^45^. A study form India reported 83.4% prevalence of low HDL cholesterol among treatment-naïve HIV-infected children aged 2 to 12 years; 52% and 58% of the children were stunted and underweight, respectively^46^. The fact that the prevalence of low HDL in our cohort is higher and consistent with that found in young adults and adults living with HIV is of concern. Some of the components of MetS acquired early in life are associated with the development of atherosclerotic lesions in children and adolescents^27,47^. HDL plays an important role in the first step of the reverse cholesterol transport pathway by removing cholesterol from cells and has anti-thrombotic and anti-atherogenic properties^48,49^. In a study from the Eastern Cape Province of South Africa in adults living with HIV, low HDL was independently associated with insulin resistance (IR) (Ref, Awotedu et al. South Africa). Low HDL is associated with inflammation resulting in IR and other cardiometabolic disorders (Ref). IR is proposed as one of the main underlying mechanisms for the development of MetS^50^. Is low HDL, therefore, a harbinger for development of MetS in these children?

The prevalence of insulin resistance in HIV+ children varies from 2% to 52% depending on age, Tanner stage, ethnicity/race, country of study, sample size, methods, and cut off for measuring insulin resistance^20,33,34,51-55^. Therefore, the prevalence of insulin resistance in our study participants is consistent with that of previous studies. The prevalence turns to be lower in prepubertal HIV+ children compared to pubertal and post pubertal HIV+ children. A study among prepubertal South African HIV+ children on ART reported insulin resistance prevalence of 2% using the HOMA-IR^17^, while Innes et al. found a much higher prevalence (10%) among prepubertal South African HIV+ children^41^. Although limited in number, longitudinal studies have uniformly found an increase in insulin resistance among CALWH with time^21,43,56^. Etiology of insulin resistance in CALWH is multifactorial including HIV infection itself, ART exposure, mitochondrial dysfunction and traditional CVD risk factors (e.g., genetic factors, nutrition, physical activity)^57-62^. Insulin resistance increases the lifetime risk of developing type 2 diabetes, CVD, and other end organ disorders^63,64^. With the high prevalence of insulin resistance with CALWH, there is the need for urgent longitudinal studies to well characterize the prevalence, risk factors, and natural history of insulin resistance in CALWH.

The main strengths of the current study are one of the first studies in sub-Saharan Africa to report on the prevalence of MetS in CALWH, raising the concern of high prevalence of components of MetS diagnostic criteria, and the random sampling design that hints that the true prevalence of MetS is high in CALWH in Uganda. However, several limitations of our study must be considered in extrapolating or interpretation of our findings to other populations. First, the lack of HIV-negative control group to determine the prevalence of components of MetS in the general population where these children reside. Second, the low numbers of MetS limited the assessment of associations of demographic and HIV disease characteristics with development of MetS. Third, lack of local population reference ranges for HOMA and lipids. Fourth, lack of nutritional history to validate the low weight and BMI in our study participants. Of note, there was no significant correlation between dietary variables and lipid or HOMA index in a study of HIV-infected prepubertal South African children on ART^41^. These limitations should be considered in designing urgent and needed studies of MetS in CALWH in sub-Saharan Africa.

## Conclusion

The high prevalence of components of MetS, particularly low HDL and early insulin resistance, are of concern. With increasing survival of CALWH into adulthood and increased lifetime exposure to ART, the frequency of MetS in this population may rise, increasing the lifetime risk for associated health problems, such as type 2 diabetes, myocardial infarction, stroke, and nonalcoholic fatty liver disease. Thus, there is a need to study the natural history of MetS in CALWH to inform preventative and treatment interventions as needed.

## Data Availability

All data produced in the present study are available upon reasonable request to the authors

## Acknowledgements

We thank the children and the families receiving care at Baylor-Uganda Clinical COE who agreed to be part of the study. We are grateful to the staff at the clinic for their support and to Elliott Paintsil for his technical assistance.

## Notes

### Competing Interest Statement

The authors have declared no competing interest.

### Funding Statement

This study did not receive any funding

### Author Declarations

Ethics committees/IRBs of School of Biomedical Sciences Higher Degree, Makerere University, Uganda, Uganda National Counsel for Science and Technology, and Yale School of Medicine gave ethical approval for this work

